# Ecological momentary assessment and mediation analyses reveal relationships between affect, craving, and substance use in daily life

**DOI:** 10.1101/2025.08.08.25333320

**Authors:** Emmanuelle Baillet, Nicholas R. Harp, Hedy Kober, Corey Roos

**Author notes:** These authors contributed equally (Hedy Kober, Corey Roos).

## Abstract

**Background and Aims:** Craving, the desire to use substances, is a key factor and diagnostic criterion for substance use disorders (SUDs) that predicts substance use. Experiencing affective states, both positive (PA) and negative (NA), are also implicated in SUDs. Yet, the interrelationships among affect, craving, and substance use remain unclear, particularly at the within-person level of analysis. This study aimed to use ecological momentary assessment (EMA) to test whether craving mediates the association between PA/NA and subsequent substance use.

**Design:** Observational study using EMA over a 14-day period and multilevel mediation models with bootstrapping.

**Setting:** Outpatient addiction treatment centers in which participants were enrolled during the data collection period.

**Participants:** Adults with SUDs enrolled in a pilot randomized controlled trial, evaluating an app-delivered mindfulness-based intervention for SUDs, that completed the baseline EMA period and were also randomized to a treatment condition (N=36).

**Measurements:** Participants completed EMA surveys three times daily [midday, afternoon, evening] for 14 days, reporting on current PA, NA, craving, and substance use. We used multilevel mediation modeling to test within-person associations and examine whether craving mediated the link between affect (PA and NA) and subsequent substance use.

**Findings:** Mediation analysis showed an indirect pathway via craving on the link between affect and substance use. Higher PA was associated with lower craving at the same EMA survey, which in turn led to reduced likelihood of substance use at the next survey (β=-0.04, p=0.02). Conversely, higher NA was associated with higher craving at the same survey, leading to greater likelihood of substance use at the next survey (β=0.08, p=0.03).

**Conclusion:** At the within-person level, craving mediates the association between affective states and subsequent substance use in individuals with SUD. PA predicted *lower* craving, which in turn predicted *lower* likelihood of substance use, while NA predicted *higher* craving, which in turn predicted *greater* likelihood of substance use. Our findings suggest that a promising strategy may be targeting momentary affect in daily life, such as through ecological momentary interventions.

## INTRODUCTION

Substance Use Disorders (SUDs) are chronic, relapsing conditions characterized by a loss of control over substance use (1, 2). Craving, a “strong desire or urge to use,” is the most central, discriminating, and prevalent criterion among all DSM-5 criteria (3-5), and across SUDs. Further, a recent meta-analysis (N=51,788) confirmed that craving prospectively predicts substance use and relapse (6), making it an important trigger of use, especially during abstinence and after recovery, even in the long-term (6-8).

Understanding antecedents of craving is critical for informing targeted interventions. A variety of antecedents – both external (e.g., seeing substances) and internal (e.g., affective states) – can elicit craving through conditioning processes (9-11). External cues are known to trigger cue reactivity, including cue-induced craving (9). One key internal cue is affect, often conceptualized along two dimensions: positive affect (PA) (e.g., joy, excitement, etc.) and negative affect (NA) (e.g., sadness, anger, etc). Several studies have reported a positive relationship between NA and craving (12, 13), summarized in a meta-analysis (N=2,257; 14). Another meta-analyses recently demonstrated that inducing NA in the laboratory leads to alcohol craving (N=2,403; 15) and nicotine craving (N=1,412; 16). Stress, a specific form of NA, has been strongly linked to craving and substance use across clinical, laboratory, and neurobiological studies (17, 18). Exposure to stressors is known to precipitate relapse (17, 18) and exacerbate withdrawal symptoms and craving, making abstinence more challenging (17, 19). This work underscores the critical role of stress and NA in triggering craving and maintaining SUDs.

Compared to NA, PA has been relatively understudied, and findings are mixed regarding its association with craving. Some studies suggest that PA reduces craving (13, 20, 21), while others suggest no direct effect (22, 23), or an indirect effect via emotion regulation strategies (24). One study utilized ecological momentary assessment (EMA) and found that momentary PA was positively associated with craving to drink among young adult heavy drinkers (25). PA may also enhance the capacity for emotion regulation and redirect the drive for rewards, thereby reducing craving (1, 26, 27). It is also possible that the effects of PA on craving depend on the subpopulation: young adult heavy drinkers are often characterized by reward drinking tendencies, and PA may be a trigger to drink (28). Alternatively, older individuals with SUDs are often characterized by relief drinking tendencies and PA may be protective (or low PA may be a trigger to use; 29).

Notably, a limitation of the work on PA/NA and craving is that it has mostly examined their association at the between-person level, and at a single timepoint, with few exceptions (e.g., 25). Testing the *within-person* association between affect and craving can reveal whether momentary changes in affect are related to changes in craving, for a given individual. Such work can reveal dynamic momentary processes occurring within individuals across time.

Beyond their effect on craving, PA and NA have been identified as potential mechanisms underlying the maintenance of substance use (30). Meta-analyses have shown that laboratory-induced NA increases substance use (15, 31), and that NA is a significant predictor of relapse (32). Another mega-analysis from 69 daily-diary and EMA studies (N=12,394) found that high NA did not predict alcohol use, whereas high PA did (33). These results suggest that PA and NA may influence substance use through different mechanisms, and potentially in different contexts.

Given the links between affect, craving, and substance use, one possibility is that craving might mediate the relationship between affect and substance use. In other words, affect may indirectly impact substance use by triggering craving. Examining this mediational pathway is crucial for understanding how affect might drive substance use. To our knowledge, two studies tested such a mediational pathway. Firstly, among patients with chronic pain using prescription opioids, craving mediated the link between NA and opioid misuse (34). Secondly, among individuals with SUDs, craving mediated the link between depressive symptoms and substance use (35). However, in (34), participants were not diagnosed with SUD, and in (35), only depressive symptoms were evaluated (not affect). Finally, these studies relied on retrospective lab-based questionnaires, limiting ecological validity and introducing potential recall bias.

Since craving and affect are dynamic fluctuating states influenced by internal and environmental factors (36, 37), EMA offers a more sensitive approach by capturing real-time states in the natural environment, through repeated measurements (38, 39). To date, no studies have investigated whether craving mediates the link between affect and substance use in daily life using EMA. The objective of this study was to use EMA to investigate the influence of affect (PA and NA) at one timepoint (T_0_) on substance use at the subsequent one (T_1_), as well as the presence of indirect effects via craving. We hypothesized that (1) PA and NA at T_0_ predict craving at T_0_; (2) craving at T_0_ predicts substance use at T_1_; and (3) craving at T_0_ would mediate that the effect of PA and NA at T_0_ on substance use T_1_.

## METHOD

### Participants

Participants were 36 individuals enrolled in a pilot randomized controlled trial (described below) evaluating an app-delivered mindfulness-based intervention for SUDs (40). Inclusion criteria were: 18 years of age or older; English-speaking; SUD diagnosis; in an early phase of SUD treatment, as demonstrated by completing at least one month of SUD treatment in the past 4 months, and using their primary substance of choice in the past 6 months; not currently enrolled in residential treatment; willingness to be randomized; willingness and ability to participate for the entire 18-week study period; willingness to provide locator information for follow-up; and own a working, WIFI-enabled smartphone. Exclusion criteria were: current psychotic symptoms; high suicide risk; homicidal ideation posing imminent danger to others; pending legal case, imminent incarceration, or a planned move that results in inability to commit to procedures during the entire study period; and participation in the previously-conducted beta-testing study (41). The protocol was approved by Yale University’s Institutional Review Board, and all participants provided informed consent.

### Study design

Following a baseline assessment visit, participants in this trial completed 2 weeks of EMA prior to being randomized (details on this trial are described elsewhere (40)). EMA consisted of 4 brief surveys per day over 2-weeks: a survey delivered in the morning, during which participants reported their past-day substance use (the “past day report” which is not used in the current analyses), and 3 additional surveys delivered randomly (every ≈4 hours) in the following time windows: midday report [12-1:30pm]; afternoon report [4-5:30pm]; evening report [8-9:30pm]. Each EMA survey was administered via the Catalyst smartphone app (Metric Wire, Inc.), which delivered smartphone notifications to complete the brief surveys. For the midday, afternoon, and evening reports, participants had 1 hour to complete them with reminders every 15 minutes. For the current analyses, we included participants who completed the baseline pre-randomization EMA period.

### Measurements

At baseline, SUD diagnosis was assessed using the Mini International Neuropsychiatric Interview (MINI; (42)). For each EMA survey (midday, afternoon, and evening), participants rated their positive affect (PA) and negative affect (NA). For example, they were asked to rate “right now (or in the past 15 minutes), I feel happy.” Participants also rated their craving (e.g., “Right now (or in the past 15 minutes), I crave, want, or desire alcohol or drugs”). Items were rated on a 10-point scale (1 not at all to 10 extremely). The PA items were: excited, grateful, sense of connection, happy, relaxed, sense of meaning/purpose; the NA items were: sad, angry, lonely, ashamed, anxious, bored. The PA/NA items were chosen based on the circumplex model of affect (43). We averaged responses within each category to calculate PA and NA scores for each random-prompt entry (i.e., average NA and PA score for the midday, afternoon, and evening surveys). At each random prompt, participants also reported substance use via two items assessing alcohol (“Did you drink any alcohol?”) or other drug use (“Have you used any other drugs, such as marijuana, cocaine, heroin, opioids, benzos, amphetamines, hallucinogens, or dissociative drugs?”). A composite binary variable indicated whether any substance use occurred. Time windows varied by survey: for midday (random prompt between noon and 1:30pm), “Since waking up today until now”; afternoon (4:00 and 5:30pm), “Since about 2pm”; and evening (8:00 and 9:30pm), “Since about 6pm”. Importantly, given the nature of time-lagged analyses and the structure of the EMA surveys, we only used prompts from midday, afternoon, and evening reports in the current analyses.

### Analyses

Our analytic plan was guided by our conceptualization of affect and craving as momentary processes occurring at the same time, and that may confer risk for subsequent substance use at the next timepoint. Accordingly, we examined: (1) the associations between PA/NA and craving; (2) the associations between PA/NA and substance use; (3) the associations between craving and substance use; and (3) whether craving mediated the link between PA/NA and subsequent substance use.

#### Link between affect and craving

We explored how fluctuations in affect intensity (PA and NA) relative to an individual’s mean level of PA and NA at each random prompt were associated with craving intensity at the same random-prompt timepoint (T_0_; concurrent effects). This approach isolates the impact of momentary deviations in affect from each participant’s average emotional experience. On an exploratory basis, we then explored whether the intensity of PA and NA at T_0_ influenced craving at the subsequent timepoint (T_1_; cross-lagged effects). These analyses accounted for temporal dynamics and allowed us to evaluate the predictive relationships between PA/NA and craving.

#### Link between affect and substance use

We explored how PA/NA fluctuations at T_0_ were associated with substance use at T_1_. Exploratorily, we also tested whether PA/NA at T_0_ predicted substance use at T_0_.

#### Link between craving and use

We explored how craving fluctuations at T_0_ were associated with substance use at T_1._

Exploratorily, we also tested whether craving at T_0_ predicted substance use at T_0_.

#### Relationship between affect, craving, and use

To explore our hypothesis that craving would mediate the relationship between PA/NA and substance use, we conducted multilevel mediation modeling (44). Mediation analysis is used to examine the interrelations among a predictor (X) (i.e., PA or NA), a mediating variable (M) (i.e., craving), and an outcome variable (Y) (i.e., substance use). Accordingly, we evaluated: the effect of PA or NA at T_0_ on craving at T_0_ (path *a*, Model X → M); the effects of craving at one time point T_0_ on substance use at T_1_ (Path *b*, Model M → Y) and then, the effect of PA or NA at T_0_ on substance use at T_1_ (Model X → Y), both as a total effect (Path *c*, without adjusting for craving) and as direct effect (Path *c’*, adjusting for craving to account for the mediating pathway). From these models, we derived the total, direct, and indirect effects. The indirect effect was computed as the product of the indirect paths (a × b). The direct effect represents the portion of PA/NA’s influence on substance use that is not mediated by craving. The indirect effect represents the portion of PA/NA’s effect on substance use that is mediated by craving. The total effect represents the overall influence of PA or NA on substance use, combining both the direct (unmediated) and indirect (mediated through craving) effects. Finally, to test the robustness of these effects, we applied bootstrapping with 1000 iterations (45), recommended for estimating the precision of effects (46). We obtained 95% confidence intervals (CI) for the direct, indirect, and total effects from the bootstrap results. To determine whether the effects were statistically significant, we computed a two-tailed p-value from the bootstrap estimates. This mediation analysis was conducted and reported in accordance with the AGReMA Statement (47).

Given the repeated and nested nature of the EMA data, we used linear mixed-effects models. For all models, we included random intercepts to account for individual differences in baseline outcome and random slopes to capture variability in the relationships between affect and craving or substance use across participants. Fixed effects were estimated to assess the associations between predictors (PA, NA, or craving) and the outcomes (craving or substance use). Models were estimated using maximum likelihood estimation with robust standard errors, and missing data were handled using full information maximum likelihood (FIML). A linear mixed model was used for PA, NA, and craving due to their ordinal nature and generalized linear mixed effects model for substance use due to its binary nature. Random intercept models were controlled for potential effects of age and sex. If a significant effect was found, it was included in the subsequent analyses. Lagged analyses controlled for prior craving or substance use, depending on the outcome. Statistical analyses were conducted in R (version 4.1.1) using the lme4 and glmer package, and code will be provided upon request.

## RESULTS

### Sample characteristics

On average, participants (N=36) were 39.5 years old (SD=9.89), 66.7% identified as men (n=24), 30.6% as women (n=11), and 2.8% as transgender (n=1; Table 1). The main primary substances reported by participants were cocaine (33.3%, n=12) and alcohol (27.8%, n=10; Table 1).

**Table 1.** Descriptive characteristics about the study sample (N=36)

| **VARIABLES** | **N (%)** | **M (SD)** |
| --- | --- | --- |
| **Sex** |  |  |
| Male | 23 (63.9) |  |
| Female | 13 (26.1) |  |
| **Gender** |  |  |
| Men | 24 (66.7) |  |
| Women | 11 (30.6) |  |
| Transgender | 1 (2.8) |  |
| **Ethnicity** |  |  |
| Hispanic | 12 (33.3) |  |
| No Hispanic | 24 (66.7) |  |
| **Race** |  |  |
| African American/Black | 13 (36.1) |  |
| White | 13 (36.1) |  |
| Other | 8 (22.2) |  |
| Prefer not to say | 2 (5.6) |  |
| **Age (years)** | 39.5 (9.89) |  |
| **Education** |  |  |
| Graduate degree | 1 (2.8) |  |
| Undergraduate degree | 8 (22.2) |  |
| Lower than secondary education | 4 (11.1) |  |
| High school diploma | 16 (44.4) |  |
| Some post-secondary education | 5 (13.9) |  |
| Other | 2 (5.6) |  |
| **Occupation** |  |  |
| Full time employment | 12 (33.3) |  |
| Part time employment | 5 (13.9) |  |
| Unemployed | 15 (41.7) |  |
| Unable to work | 3 (8.2) |  |
| Student | 1 (2.8) |  |
| **Primary substance** |  |  |
| *Cocaine* | 12 (33.3) |  |
| *Alcohol* | 10 (27.8) |  |
| *Dissociative drug (PCP, ketamine)* | 6 (16.7) |  |
| *Marijuana* | 4 (11.1) |  |
| *Heroin* | 2 (5.56) |  |
| *Hallucinogens (MDMA, LSD)* | 1 (2.78) |  |
| *Opioid pills (not prescribed or misused)* | 1 (2.78) |  |
| **DSM-5 SUD** |  |  |
| *Sum of criteria (0-11)* |  | 8.5 (2.9) |
| *Mild (2-3)* | 4 (11.1) |  |
| *Moderate (4-5)* | 4 (11.1) |  |
| *Severe (6+)* | 28 (77.8) |  |
| **Treatment & Medication** |  |  |
| SUD medication | 11 (30.5) |  |
| Previous inpatient/residential treatment episodes for SUD |  | 2.85 (4.05) |
| Previous outpatient treatment episodes for SUD |  | 3.86 (3.89) |
| Referred by the criminal justice system | 27 (75%) |  |
| **EMA features** |  |  |
| Completed surveys | 1,331 (66.1)<sup>1</sup> |  |
| Craving (1-10) |  | 1.91 (2.26, range 1-10) |
| Positive Affect (1-10) |  | 6.0 (2.35, range 0-10) |
| Negative Affect (1-10) |  | 2.5 (1.77, range 0-8.6) |
| Substance use | 193 (21.44) |  |

**Note.** M = mean; SD = standard deviation.
<sup>1</sup>Percentage calculated out of the maximum possible number of EMA surveys.

### EMA completion

With four EMAs per day over fourteen days, each participant received 56 prompts, resulting in a potential maximum of 2,016 completed EMAs (n= 56×36). Of this, there were 665 missed EMAs (32.9%) and 1,331 completed EMAs (67.1%), which is an acceptable compliance threshold (48). Of the four EMAs per day, percent completion ranged from 85.5% for the first EMA of the day to 57.9% for the last EMA of the day.

### Affect, craving, and substance use

Overall, the average score was 2.16 (SD=1.42, range=0-10) for craving, 6.03 (SD=1.19, range=0-10) for PA, and 2.47 (SD=0.98, range=0-8.6) for NA. Substance use was reported at 20.5% (SD=28.5) of the midday EMA surveys, 20.9% (SD=28.5) of the afternoon surveys, and 25.7% (SD=30.7) of the evening surveys.

### Link between affect and craving

Prior to model estimation, craving variance during the EMA study was explored using intraclass correlation (ICC). An ICC value of 0.62 indicated that craving variance was attributed to between-person differences (61.7%) and within-persons differences (38.3%). Sex and age did not predict craving (Sex: β=-0.09; SE=0.85; χ^2^=-0.11; p=0.91; Age: β=-0.01; SE=0.04; χ^2^=-0.26; p=0.80) and thus, were not included in the following analyses. Multilevel models revealed that both PA and NA at T_0_ predicted craving at T_0_. However, their effects differed: higher levels of PA were associated with decreased craving intensity (β=-0.23; SE=0.05; χ^2^=-4.32; p<0.001), whereas higher levels of NA were associated with increased craving intensity (β=0.55; SE=0.05; χ^2^=10.21; p<0.001; Figure 1). Exploratory analyses revealed that NA at T_0_ predicted craving at T_1_ (β=0.29; SE=0.29; χ^2^=3.62, p<0.001; Figure 2), but this was inconclusive for PA (β=-0.01; SE=0.06; χ^2^=-0.24, p=0.808).

**Figure 1:**
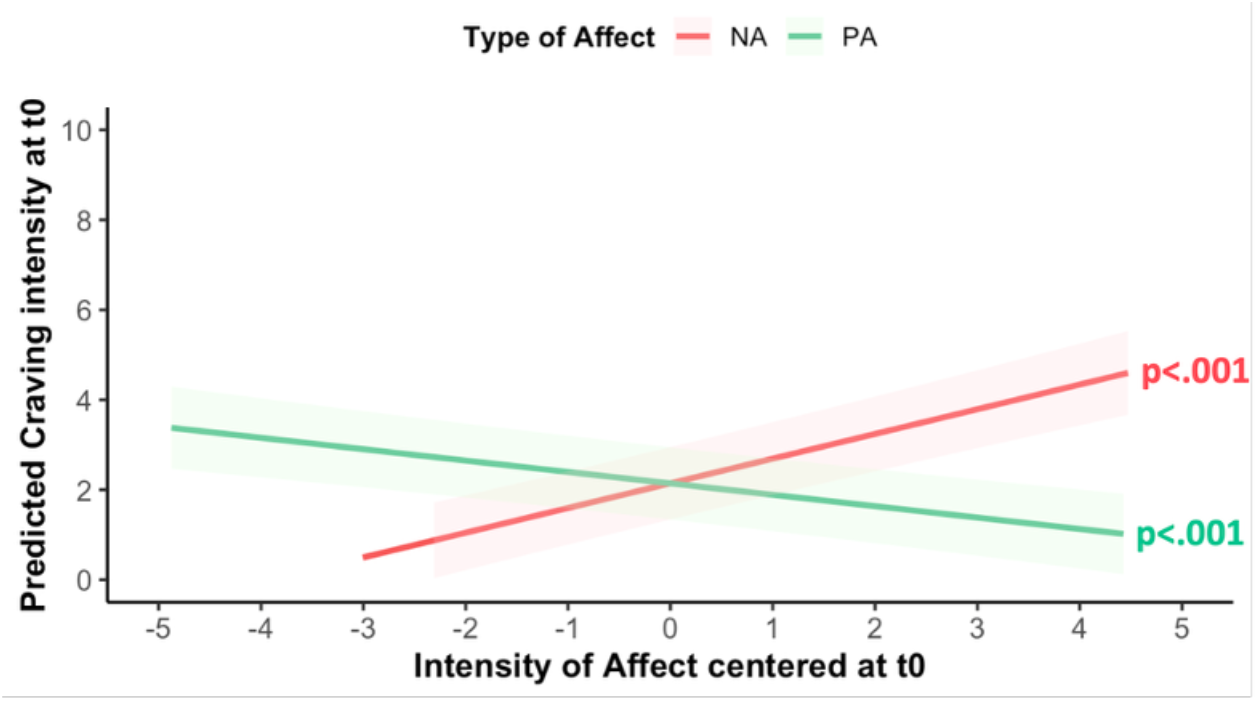
Predicted effects of positive (green) and negative (orange) affect at one EMA timepoint (T_0_) on craving intensity at the same one (T_0_). Shaded areas indicate 95% confidence intervals (fixed effects, centered affect variables).

**Figure 2:**
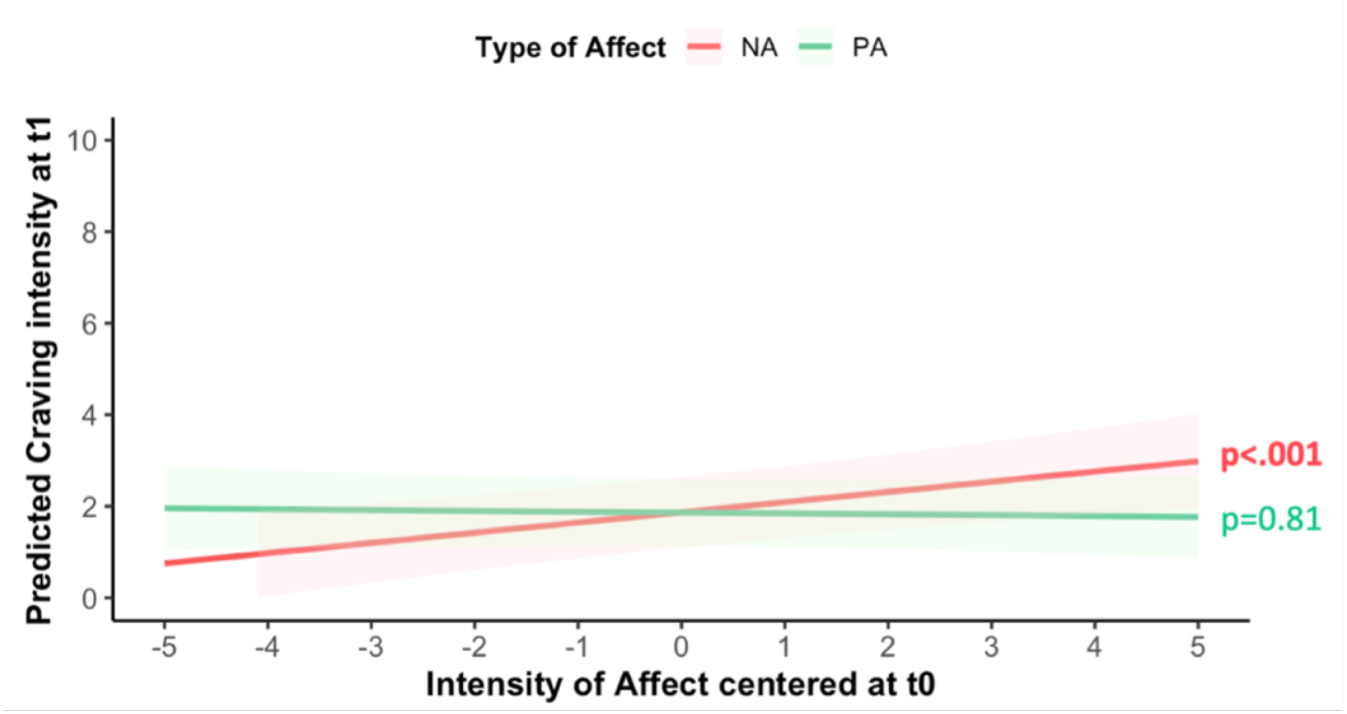
Predicted effects of positive (green) and negative (orange) affect at one EMA timepoint (T_0_) on craving intensity at the next timepoint (T_1_). Shaded areas indicate 95% confidence intervals (fixed effects, centered affect variables, controlled for craving at T_0_).

### Link between affect and use

Substance use had an ICC value of 0.53, showing between-person variation accounted for 78.4 % of the variance, while within-person variation accounted for 21.6%. Sex and age did not predict substance use (Sex: β=-1.06; SE=0.74; χ^2^=-1.44; p=0.15; Age: β=-0.01; SE=0.04; χ^2^=-0.31; p=0.76) and were not included in the following analyses. There were inconclusive effects of PA and NA at T_0_ on substance use at T_1_ (PA: β=-0.01; SE=0.09; χ^2^=-0.07, p=0.941; NA: β=0.17; SE=0.12; χ^2^=1.43, p=0.152). Similarly, exploratory analyses showed inconclusive effect of either PA or NA at T_0_ on substance use at T_0_ (PA: β=-0.02; SE=0.07; χ^2^=-0.306, p=0.76; NA: β=0.17; SE=0.09; χ^2^=1.81, p=0.07).

### Link between craving and use

Craving at T_0_ predicted substance use at T_1_, after controlling for substance use at T_0_ (β=0.14; SE=0.07; χ^2^=2.06; p=0.04; Figure 3). Exploratory analyses showed that craving also predicted substance use at T_0_ (β=0.15; SE=0.05; χ^2^=2.69; p=0.01; Figure 4).

**Figure 3:**
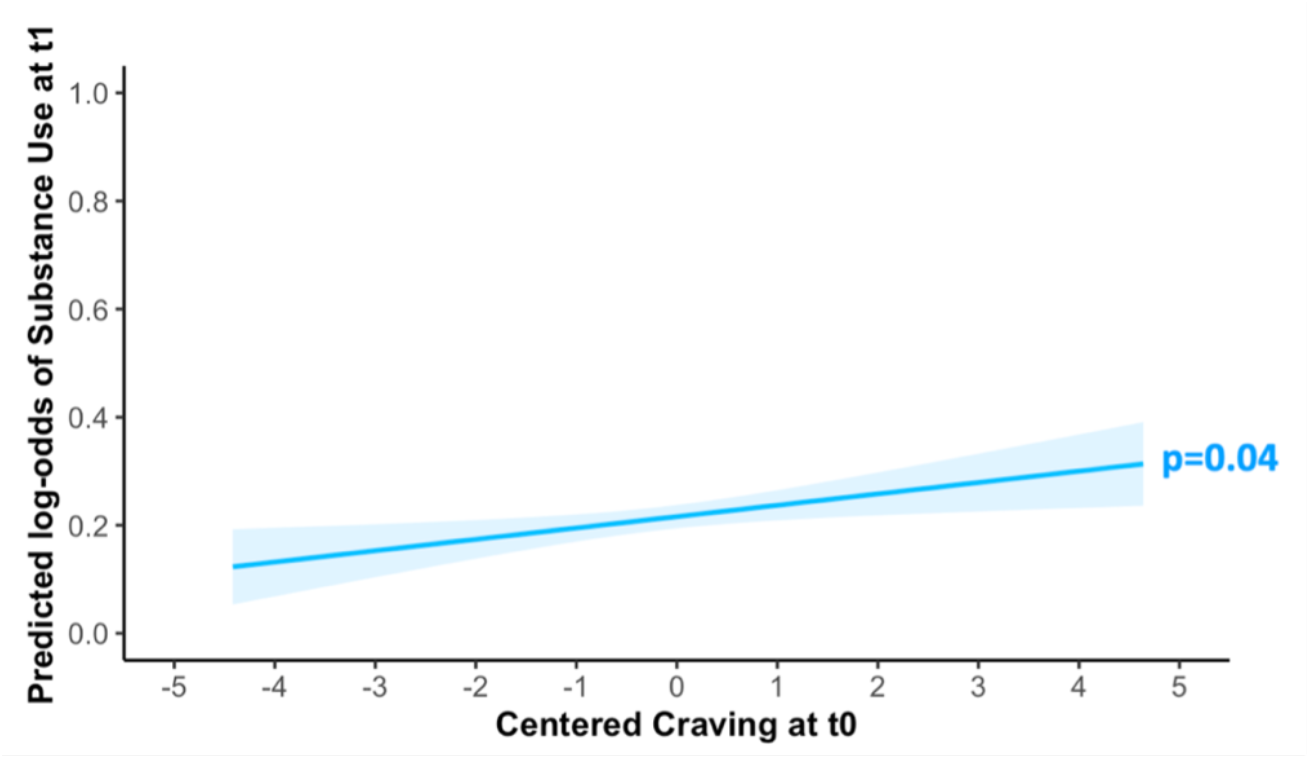
Predicted effects of craving at one EMA timepoint (T_0_) on the substance use at the next timepoint (T_1_). Shaded area indicates 95% confidence interval (fixed effect, centered craving variable, controlled for substance use at T_0_).

**Figure 4:**
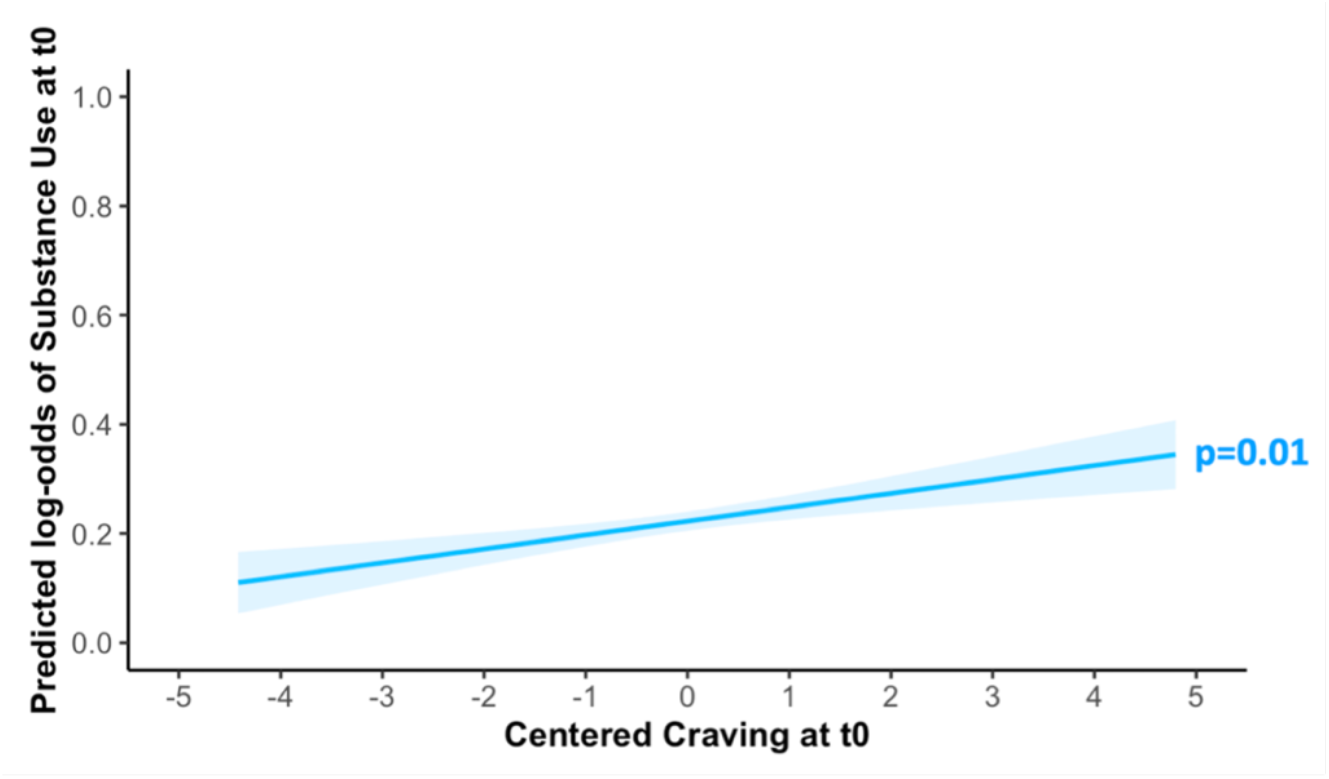
Predicted effects of craving at one EMA timepoint (T_0_) on the substance use at the same one (T_0_). Shaded area indicates 95% confidence interval (fixed effect, centered craving variable).

### Relationship between affect, craving, and use

The total effect of PA and NA at T_0_ on substance use at T_1_ was not significant (PA: β=-0.01, SE=0.11, 95% CI [-0.215, 0.217], p=0.96; NA: β=0.18, SE=0.12, 95% CI [-0.078, 0.429], p=0.15; Figure 4). Controlling for craving did not change this result (direct effect/PA: β=0.03, SE=0.11, 95% CI [-0.187, 0.256], p=0.82; NA: β=0.10, SE=0.13, 95% CI [-0.144, 0.364], p=0.42; Figure 5). This indicates that neither PA nor NA directly predict substance use over time. However, our results revealed significant indirect effects between PA/NA at T_0_ and substance use at T_1_, such that the effect of PA or NA at T_0_ on substance use at T_1_ was mediated by craving at T_0_ (indirect effect for PA: β=-0.04, SE=0.02, 95% CI [-0.087, -0.002], p=0.02; for NA: β=0.08, SE=0.04, 95% CI [0.006, 0.164], p=0.03; Figure 5). Specifically, the negative indirect effect of PA suggests that higher PA is associated with lower craving, which in turn leads to reduced substance use. Conversely, the positive indirect effect of NA indicates that higher NA is linked to increased craving, ultimately leading to greater substance use. These findings indicate that the relationship between PA/NA and substance use is primarily explained through craving rather than a direct influence of PA or NA on substance use.

**Figure 5:**
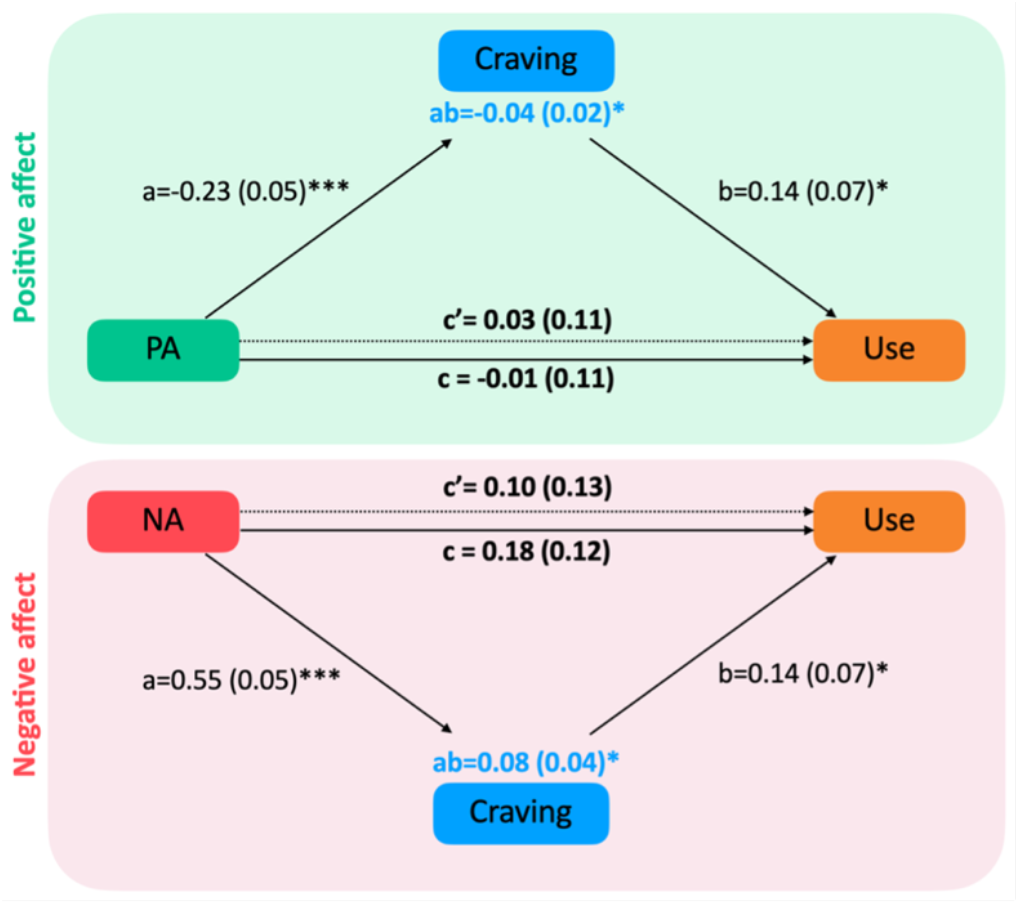
Mediation models for the association between affect (PA or NA) at one EMA timepoint (T_0_), craving at the same timepoint (T_0_), and substance use at the next timepoint (T_1_), whereby craving is a complete mediator of the affect–use relationship. Path coefficients are presented alongside arrows representing each analyzed link, with standard errors (SEs) in parentheses. Path *a* denotes the effect of affect on craving, while path *b* represents the effect of craving on substance use. Path *c’* illustrates the relationship between affect and substance use while accounting for the mediator (craving), whereas path *c* represents the total association between affect and substance use without considering the mediator. Significance levels are indicated as follows: *p<0.05, **p<0.01, ***p<0.001.

## DISCUSSION

This EMA study examined how momentary affect, craving, and substance use interact in daily life among individuals with SUDs. Results confirm that craving mediates the association between affect and substance use: PA predicted *lower* craving, which in turn predicted *lower* likelihood of substance use. while NA predicted *higher* craving, which in turn predicted *greater* likelihood of substance use. These findings confirm the key role of affect and craving in the dynamics of substance use.

Our findings are consistent with prior research showing that NA predicts craving; however, here we show that this association also occurs at the within-person level. The findings are also consistent with neurobiological models of SUDs that have highlighted the role of stress and NA in craving and SUDs (2). Prior work on the association between PA and craving has primarily examined effects at the between-person level, and results have been mixed. Here, we show that PA was associated with lower craving and may thus protect against craving at the within-person level, in line with some prior work (49, 50). High PA may correspond with non-substance-related rewarding experiences in daily life. Hence, individuals may be less tempted to use substances because they are experiencing alternative rewards (51, 52).

Importantly, we found that affect did not predict substance use at T_0_ or at T_1_. Rather, our results highlight an indirect pathway: PA was associated with reduced craving, which in turn was associated with lower subsequent substance use, while NA was associated with increased craving and substance use. This suggest that affect influences substance use indirectly via its effect on craving. Notably, these results conflict with some prior reports of a direct positive link between PA and alcohol use (33). This discrepancy may be due to differences in sample characteristics (multi-substances vs. alcohol only), temporal resolution (momentary vs. daily-level data), and analytic approach (craving not directly modeled in analyses). Additional research is needed to elucidate the conditions under which PA may affect craving and use across time resolutions, samples, etc.

Exploratory analyses showed NA increases craving at both at T_0_ and T_1_, while PA reduces craving only at T_0_. These aligns with previous studies demonstrating that momentary affect can trigger craving (53-56). The novel temporal associations suggest that NA may have a more persistent impact on craving, beyond the moment when it is experienced. Conversely, PA could be more short-lived, offering only a temporary distraction from craving. Notably, these findings further highlight the temporal nature of these relationships in daily life, while highlighting the critical role of craving in drug use.

Importantly, the mediation analysis suggests that associations between affect and subsequent substance use are indirect, operating through craving. Neither PA nor NA directly predicted substance use. However, both significantly influenced substance use via their momentary association with craving. It is worth noting that the presence of significant indirect effects in the absence of total or direct effects may appear counterintuitive. However, such a pattern has been well documented in the literature (57, 58), and can occur when multiple opposing pathways cancel each other out, or when indirect paths are simply more sensitive to detecting subtle effects. Overall, these findings emphasize craving as a key central mechanism connecting momentary affect to substance use in everyday life. It also supports influential addiction theories suggesting that craving is a proximal predictor of use influenced by emotion states (6, 17, 18, 59, 60).

This study highlights the potential of interventions targeting daily affect to reduce substance use via craving. Just-in-time adaptive interventions (JITAIs), such as mindfulness or cognitive behavioral treatments, may help individuals regulate NA, or sustain PA (61, 62), indirectly reducing craving and subsequent use (63), along with strategies that directly modulate craving (64). This study has strengths including real-time EMA data capturing within-day dynamics and identifying the mediating role of craving. Limitations include possible unmeasured fluctuations and a relatively small and clinically severe sample; future investigations should test whether these findings generalize to less diverse populations in term of severity and type of substance. Finally, the mediation model was based on data from an observational design which limits causal interpretation (65). Nevertheless, we believe that these results are novel, interesting, and informative about the dynamics of affect, craving, and substance use in daily life.

## Data Availability

All data produced in the present study are available upon reasonable request to the authors

## Acknowledgements

We thank all the participants who took part in this study and the research staff who contributed to data collection and management. We are grateful to the Yale University School of Medicine and the University of California, Berkeley, for institutional support. HK and CR developed the study design and methods. EB and HK conceptualized and conducted the analyses, interpreted the data, and wrote the manuscript. NH helped in analysis. All authors undertook the critical revision of the manuscript for important intellectual content and all authors significantly contributed to the manuscript and approved the final version.

## Declarations of competing interest

None.

## Primary funding

This project was supported by K23AT011342 to CR; Drs. Baillet, Harp, Kober, and Roos were also supported by R01AA029137 to HK.

## Notes

### Competing Interest Statement

The authors have declared no competing interest.

### Author Declarations

Ethics committee/IRB of Yale University gave ethical approval for this work.

